# SDF, SAP P_11_-4, and GI Sealants for Managing Initial Caries Lesions Produce Clinic and Payor Savings in Financial Impact Model

**DOI:** 10.1101/2023.01.27.23285118

**Authors:** Savyasachi V. Shah, Laura J Kibbe, Lisa J. Heaton, Courtney Desrosiers, John Wittenborn, Mariya Filipova, Kirill Zaydenman, Jeremy Horst Keeper

**Author notes:** These authors contributed equally to this work (Savyasachi V. Shah, Laura J Kibbe).

## Abstract

**Introduction:** Evidence-based noninvasive caries therapies for initial caries lesions recently became available in the United States. Fundamental differences between noninvasive therapies and the traditional surgical dental approach warrant study of the financial scalability.

**Methods:** The financial costs and benefits to fee-for-service clinics and payors were compared across eleven scenarios simulating the treatment of 1,000 initial lesions over a three-year period. The scenarios included varying combinations of noninvasive therapies (silver diamine fluoride (SDF), SAP P_11_-4, and glass ionomer sealants), no treatment, and various rates of one to three surface restorations to an estimated current practice model. We used a decision tree microsimulation model for deterministic and probabilistic sensitivity analyses. We derived assumptions from an initial lesion and noninvasive therapy-focused cohort study with operations data from 16 sites accepting Medicaid in Alabama as a case study and clinical data from all 92 sites.

**Results:** In comparison to the current practice model, scenarios that produce mutually beneficial results for payors savings and clinics net profit and profit margin include: SAP P_11_-4, SDF on non-cosmetic surfaces, and a mix of three noninvasive therapies. When considering the limited resources of chair and clinician time, the same scenarios as well as SDF with restorations emerged with substantially higher clinic net profit.

**Conclusion:** Scenarios that include noninvasive therapies and minimize restorations achieve the balance of improving outcomes for all parties.

**Practical implications:** Payors should appropriately reimburse and clinics should adopt noninvasive caries therapies to improve oral health for all.

## Introduction

Initial caries lesions are the first noticeable stage of dental caries, where a frank cavitation into dentin has not yet formed.^1^ International consensus and United States (US) guidelines state consistently that drilling and restoring are not indicated for initial lesions.^2–4^ There appears to be poor awareness that demineralization reaching as deep as the outer third of the dentin is unlikely to be cavitated and therefore infected.^1^ Historically it has been ubiquitous for dental boards to have candidates drill and restore initial lesions to obtain a dental license. Further, 94% of U.S. dentists self-report performing restorations when radiographic demineralization is still within the range of initial lesions.^5^ The same guidelines mentioned above indicate noninvasive treatments for initial lesions.^2, 3^ Tooth-specific noninvasive caries therapies include silver diamine fluoride (SDF), Self-assembling peptide (SAP P_11_-4 such as Curodont Repair), and glass ionomer sealants (sealants), among others.^3, 6, 7^ Noninvasive therapies have similar clinical outcomes to more invasive treatments without irreversibly removing tooth structures, are completed quickly and painlessly, and are often reimbursed by payors (e.g., Medicaid). However, to our knowledge, the clinic net profit and payor savings of noninvasive therapies compared to predominant current practice have not yet been studied.^4, 5^

SDF is a brush-on therapy that prevents and arrests caries lesions at any stage excluding irreversible pulpitis. SDF-treated lesions turn black, while healthy enamel stays white.^8, 9^ There have not been sufficient clinical trials to assess the efficacy of SDF on initial lesions.^10^ However, numerous clinical trials^3, 10^ support the American Dental Association (ADA)’s recommendation to arrest cavitated caries lesions on coronal surfaces using SDF. As initial lesions can be arrested with less-potent agents (e.g., fluoride varnish), it is presumed that SDF, a more potent agent, will readily arrest initial lesions.^3, 10^

SAP P_11_-4 is a biomimetic self-assembling peptide professionally applied as a brush-on liquid. SAP P_11_-4 regenerates hydroxyapatite within the porous enamel of an initial lesion by absorbing into the lesion, self-assembling into scaffolds that span throughout the demineralized volume, and templating calcium and phosphate from the saliva into new hydroxyapatite.^11^ SAP P_11_-4 has clinically important effects on arresting caries and shrinking initial lesions^6, 7^ while being safe.^12^ In January 2019, SAP P_11_-4 was added to the US Food and Drug Administration’s National Drug Codes Directory under the Over-the-Counter Anticaries Drug Monograph (NDC: 72247-101).

The ADA recommends the use of sealants in both preventing caries and arresting initial lesions in pits and fissures.^3, 13, 14^ Glass ionomer can be professionally applied to create a physical barrier and strengthen the tooth with fluoride and metal ions (e.g., remineralization), which makes the tooth less dissolvable by acid and less colonizable by bacteria.^15–17^ Due to the incorporation of fluoride and metal ions, caries prevention and arrest are maintained even if the bulk of the sealant falls out.^18^

SDF, SAP P_11_-4, and sealants are evidence-based, noninvasive therapies that can be alternatives to no-treatment or restorative approaches that could increase clinic net profits and reduce payor costs while benefiting patients’ dental treatment experience. This study analyzes the clinic costs, net profit, and profit margin, as well as payor costs of using these three noninvasive therapies to treat initial lesions compared to other restoration approaches and no treatment.

## Methods

### Study Overview

Clinic costs, gross revenue, net profit, and profit margin and payor costs were modeled for eleven scenarios of different combinations for the three noninvasive therapies (SDF, SAP P_11_-4, sealants), restorations, and no treatment in 1,000 permanent teeth with initial caries lesions. For accuracy in larger scale models, numbers were not rounded to the nearest whole tooth and therefore fractions of teeth are included in the modeling. We used data from clinics in the state of Alabama that accepted Medicaid as representative of a US region with a high disease burden and limited dental and financial resources. We used a model of estimated *current practice* to which we compared other models in which 40% of teeth with initial lesions received restorative treatment while 60% received no treatment (see current practice model rationale below).

All scenarios were modeled for this purpose with assumptions derived from a cohort as described below. The decision tree microsimulation model was built in Microsoft Excel (Redmond, WA) as per guidelines from the International Society for Pharmacoepidemiology and Outcomes Research.^19, 20^ This model was used for deterministic and probabilistic sensitivity analysis across the scenarios over three years to compare the costs, net profit, and profit margin (all in US dollars) to fee-for-service clinics and the fee-for-service payor savings. A three-year period was chosen to reflect the average duration members remain with a dental insurance plan. We did not adjust for inflation as the primary objective was to compare treatment approaches for managing initial caries lesions specifically, not the total cost of managing the patient or population.

### Data Source

The cohort consists of patients seen in ninety-two Advantage Dental+ clinics in eight states (WCG IRB#: 1295572; clinicaltrials.gov ID: NCT04933331). Sixteen of these clinics are in Alabama. These clinics focus on diagnosis and non-invasive management of initial lesions. The cohort includes 260,594 initial lesions diagnosed in 76,888 patients, with up to 6 years of follow-up. All data for the current study come from the clinical management of this cohort, except where otherwise noted. Details of the model variables involving current practice, clinician cost and time, material costs, clinic gross revenue/payor costs, and treatment failures are presented below. Table 1 lists estimated labor salaries, overhead costs, material costs, reimbursements, procedure times, and ranges thereof.

**Table 1.**
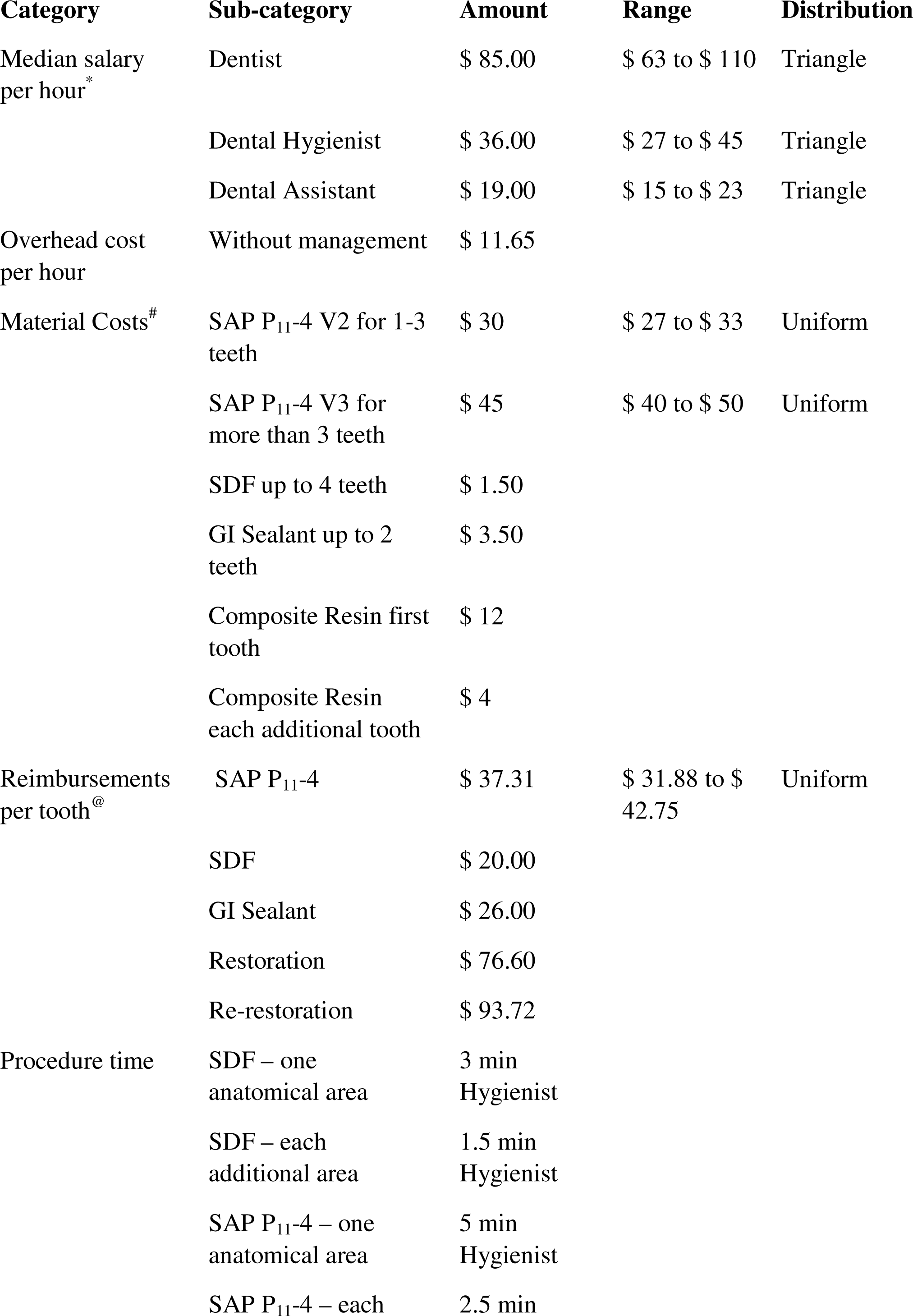
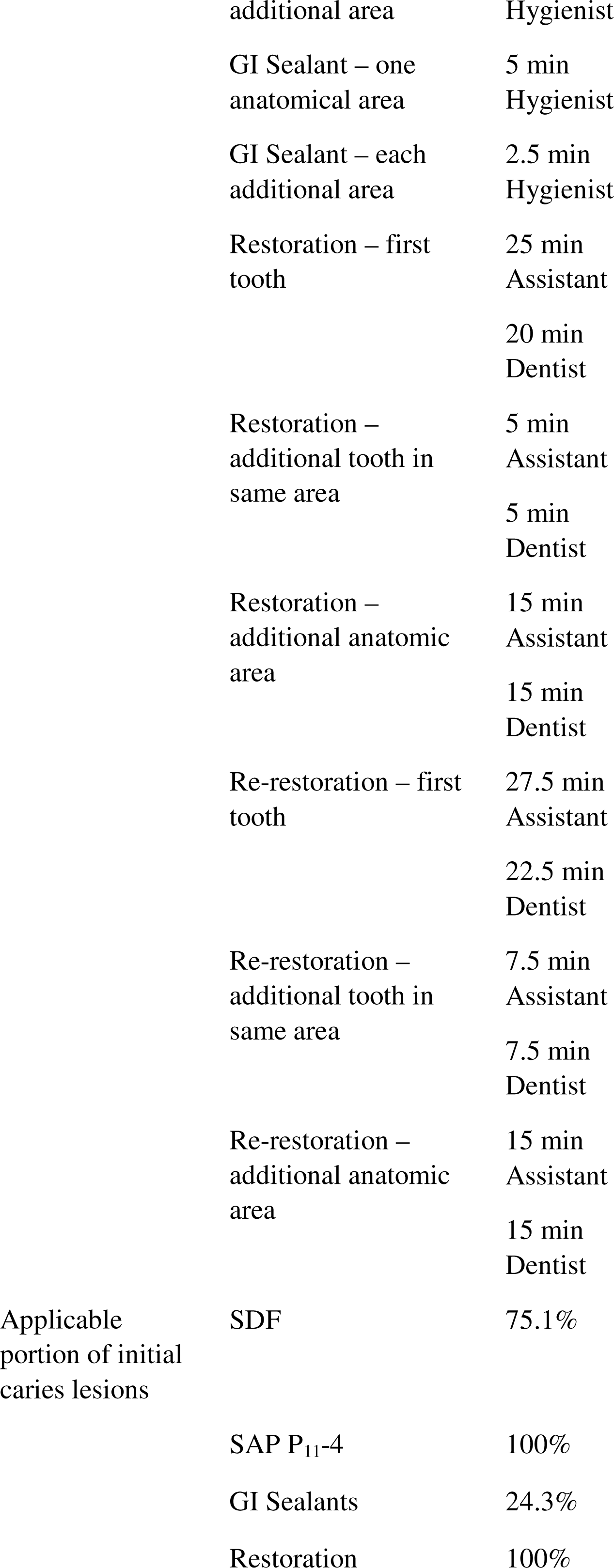
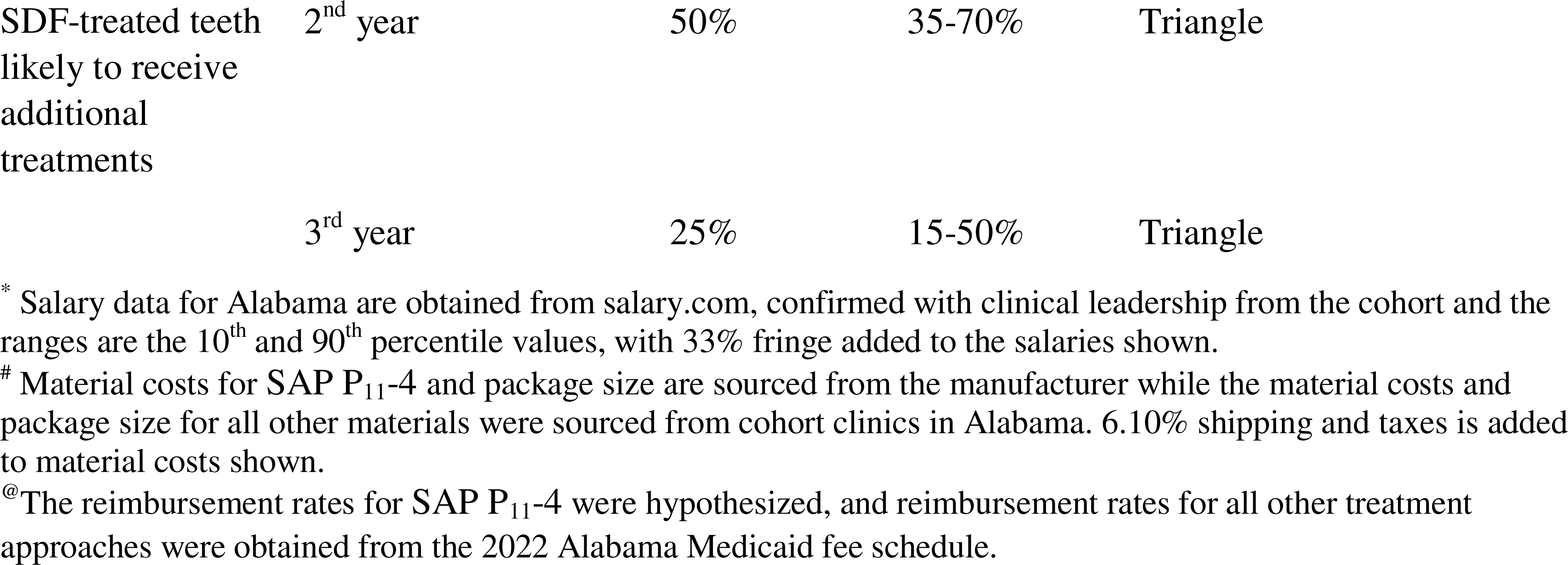
Treatment cost factor estimates: estimated hourly wages, overhead cost, material costs and reimbursements, procedure times, applicable surfaces, and ranges thereof. These data are used as input in the probability sensitivity analysis. The type of distribution for each variable, where relevant, is listed at right.

| Category | Sub-category | Amount | Range | Distribution |
| --- | --- | --- | --- | --- |
| Median salary per hour* | Dentist | \$ 85.00 | \$ 63 to \$ 110 | Triangle |
| | Dental Hygienist | \$ 36.00 | \$ 27 to \$ 45 | Triangle |
| | Dental Assistant | \$ 19.00 | \$ 15 to \$ 23 | Triangle |
| Overhead cost per hour | Without management | \$ 11.65 | | |
| Material Costs<sup>#</sup> | SAP P<sub>11</sub>-4 V2 for 1-3 teeth | \$ 30 | \$ 27 to \$ 33 | Uniform |
| | SAP P<sub>11</sub>-4 V3 for more than 3 teeth | \$ 45 | \$ 40 to \$ 50 | Uniform |
| | SDF up to 4 teeth | \$ 1.50 | | |
| | GI Sealant up to 2 teeth | \$ 3.50 | | |
| | Composite Resin first tooth | \$ 12 | | |
| | Composite Resin each additional tooth | \$ 4 | | |
| Reimbursements per tooth<sup>@</sup> | SAP P<sub>11</sub>-4 | \$ 37.31 | \$ 31.88 to \$ 42.75 | Uniform |
| | SDF | \$ 20.00 | | |
| | GI Sealant | \$ 26.00 | | |
| | Restoration | \$ 76.60 | | |
| | Re-restoration | \$ 93.72 | | |
| Procedure time | SDF – one anatomical area | 3 min Hygienist |  |  |
|  | SDF – each additional area | 1.5 min Hygienist |  |  |
|  | SAP P<sub>11</sub>-4 – one anatomical area | 5 min Hygienist |  |  |
|  | SAP P<sub>11</sub>-4 – each | 2.5 min |  |  |

|  |  |  |
| --- | --- | --- |
|  | additional area | Hygienist |
|  | GI Sealant – one anatomical area | 5 min<br>Hygienist |
|  | GI Sealant – each additional area | 2.5 min<br>Hygienist |
|  | Restoration – first tooth | 25 min<br>Assistant |
|  |  | 20 min<br>Dentist |
|  | Restoration – additional tooth in same area | 5 min<br>Assistant |
|  |  | 5 min<br>Dentist |
|  | Restoration – additional anatomic area | 15 min<br>Assistant |
|  |  | 15 min<br>Dentist |
|  | Re-restoration – first tooth | 27.5 min<br>Assistant |
|  |  | 22.5 min<br>Dentist |
|  | Re-restoration – additional tooth in same area | 7.5 min<br>Assistant |
|  |  | 7.5 min<br>Dentist |
|  | Re-restoration – additional anatomic area | 15 min<br>Assistant |
|  |  | 15 min<br>Dentist |
| Applicable portion of initial caries lesions | SDF | 75.1% |
|  | SAP P<sub>11</sub>-4 | 100% |
|  | GI Sealants | 24.3% |
|  | Restoration | 100% |
| SDF-treated teeth likely to receive additional treatments | 2<sup>nd</sup> year | 50% | 35-70% | Triangle |
|  | 3<sup>rd</sup> year | 25% | 15-50% | Triangle |

\* Salary data for Alabama are obtained from salary.com, confirmed with clinical leadership from the cohort and the ranges are the 10<sup>th</sup> and 90<sup>th</sup> percentile values, with 33% fringe added to the salaries shown.

### Current Practice Model

To determine a base case of current practice for comparison, we estimated the *expected* rate of restoration for initial lesions. Dental health records typically include treatment codes rather than diagnostic codes. Therefore, we consulted 20 experts with a national perspective on clinical care and claims data with representation from payors, nonprofit institutes, academia, and DSOs and estimated that 40% of initial lesions are treated with 1-3 surface restorations. We considered that shallower (less severe) lesions are most prevalent and least often drilled, while deeper initial lesions are nearly always drilled but are less prevalent. We verified this expected initial lesions restoration rate by cross-referencing the incidence of initial lesions and 1-3 surface restorations in the cohort data.

### Clinician Costs and Time

We estimated the clinician and chair overhead time required to perform each treatment per patient from consultation with individuals responsible for clinical management of this cohort. We then projected this estimate to reflect the quantity of teeth treated and their distribution within the four quadrants of the mouth, accounting for economies of scale when multiple lesions occur in the same patient. We modeled hygienists as providers for noninvasive therapies and dentists with an assistant for restorations. Estimates for provider time for each procedure per first and additional initial lesion(s) and quadrant(s) are shown in Table 1. No additional time was calculated per additional tooth in the same quadrant for SDF, SAP P_11_-4, or sealants. Chair time for a procedure was estimated as either assistant (restoration) or hygienist (noninvasive therapies) time. The median hourly wages for Alabama dentists, hygienists, and dental assistants were obtained from Salary.com (accessed November 25^th^, 2022) and 33% in indirect costs was added for payroll taxes and benefits. These wage estimates were confirmed to be accurate with individuals responsible for the clinical management of the cohort. The probability of the number of teeth and distribution across four anatomical areas were factored into the model to account for economies of scale in labor and material cost (Supplemental Table 1).

### Material Costs

Material costs were estimated based on unit or dose cost and the number of teeth that can be treated by each (Table 1). Cost for a planned more efficient SAP P_11_-4 product for 4+ teeth was estimated by the manufacturer. A 6.1% sales tax was added to material costs, as the average of those in the Alabama clinic locations. The total treatment costs for each intervention were estimated by considering the labor, overhead, and material costs, and treatment time, based on the number of teeth treated and their anatomic distributions observed in the cohort.

### Clinic Gross Revenue / Payor Costs

Fees were taken from the 2022 Alabama Medicaid fee schedule for each procedure code in the existing Current Dental Terminology (CDT) and are listed in Table 1. SDF was modeled with annual application with attrition, whereas SAP P_11_-4 and sealants were modeled with a single application (Table 1). The weighted average reimbursement for restorations was calculated based on the relative incidence in cohort patients for each of the nine CDT procedure codes for 1-3 surface restorations and similarly for re-restoring any overlapping surface. For the initial placement of treatment, there are economies of scale (reduced time and cost) represented when multiple lesions occur in the same patient to match clinical practice (see Supplemental Table 2). The reimbursement for restoration within one year of any noninvasive therapy was calculated by subtracting the reimbursement given for the prior treatment from the reimbursement for the restoration. No compensation is given for re-restoration within one year in Alabama. To account for multiple failures in the same tooth in sequential post-treatment years, separated by more than 12 months, 50% of such repeat failures were reimbursed in the second and third years. To establish a reimbursement rate and range for SAP P_11_-4, a full deterministic analysis was run for the current practice model and for SAP P_11_-4 with a range from $20 to $50 per tooth. The break-even for clinic net profit and payor cost with respect to the current practice model defined the minimum and maximum of the range, and the midpoint was used in further analysis in attempt to balance payor and clinic interests.

### Treatment Failure / Patient Benefit

Treatment failure is defined as any tooth/surface receiving restoration after baseline (Table 2); this affects costs. The greatest benefit to the patient is greater drill-free tooth survival (Figure 5; Supplemental Table 3), preventing the need for additional restorative visits after baseline treatment. Treatment failures were modeled at the end of each of 3 years. Due to the low treatment failure rate, we anticipated failures to be independent of any economies of scale that would occur with multiple lesions within one patient. Failure rates were derived from claims-level records from the cohort data for the proportion of teeth with operative treatment (restoration, endodontics, extraction, or crown lengthening) performed on an overlapping surface of an initial lesion previously treated with noninvasive therapies or 1-3 surface restorations. The break-even for clinic net profit and payor cost with respect to the current practice model defined the minimum and maximum of the range, and the midpoint was used in further analysis in attempt to balance payor and clinic interests.

**Table 2:** Treatment survival rates for initial caries lesions management. Per-tooth treatment failure and survival from cohort data, unless otherwise noted. These data are used to define the beta distributions for treatment failure rates in the probability sensitivity analysis. †combination of control groups in Griffin et al., 2008 and Horst Keeper et al., *in press*. *Data from previous year used due to lack of robust data. In the model, survival rates were not rounded to the nearest whole tooth and fractions of teeth are included in the modeling for greater accuracy.

| **Materials** | **Failures** | **Observations** | **Survival proportion** |
| --- | --- | --- | --- |
| **Year 1 (from 6-12 month observations)** |  |  |  |
| SDF | 400 | 15,648 | 97.4% |
| SAP P<sub>11</sub>-4 | 61 | 1,717 | 96.4% |
| Sealant | 4 | 398 | 99.0% |
| 1-3 surface restoration | 3,447 | 119,525 | 97.1% |
| Untreated† | 57 | 473 | 88.0% |
| **Year 2 (from 12-24 month observations)** |  |  |  |
| SDF | 447 | 14,579 | 96.9% |
| SAP P<sub>11</sub>-4 | 34 | 945 | 96.4% |
| Sealant | 12 | 469 | 97.4% |
| 1-3 surface restoration | 6,682 | 177,601 | 96.2% |
| Untreated†* | 57 | 473 | 88.0% |
| **Year 3 (from 24-36 month observations)** |  |  |  |
| SDF | 137 | 4,577 | 97.0% |
| SAP P<sub>11</sub>-4* | 34 | 945 | 96.4% |
| Sealant* | 5 | 229 | 97.8% |
| 1-3 surface restoration | 4,784 | 120,240 | 96.0% |
| Untreated†* | 57 | 473 | 88.0% |

### Simulation Model of Initial Caries Management

We developed 11 scenarios to reflect several different approaches to initial lesions management, and a formal model was built to reflect the 11 scenarios with 1000 teeth each (see Figure 1). In addition to one condition that involved no treatment (“NoTx”), we included four scenarios in which differing percentages of initial lesions were treated with traditional restorations while the remaining percentages remained untreated. For example, in the scenario labeled “20%Restore,” 20% of initial lesions received traditional restorations while the remaining 80% were left untreated. We created two scenarios with SDF applied on 75% of initial lesions, corresponding to the fraction of initial lesions occurring anywhere on permanent teeth except the facial surfaces of anterior and premolar teeth in the cohort data: in the “75%SDF” scenario, 75% of initial lesions were treated with SDF and 25% were left untreated; in the “SDF+Restore” scenario, 75% of initial lesions were treated with SDF and 25% were treated using traditional restorations. As sealants can only be applied to pits and fissures, similarly, we created two scenarios with sealants applied to 24% of initial lesions, corresponding to the fraction of initial lesions of permanent teeth occurring in pits and fissures in the cohort data: in the “24%Seal” scenario, 24% of initial lesions were treated with sealants and 76% were left untreated; in the “Seal+Restore” scenario, 24% of initial lesions were treated with sealants and 76% were treated using traditional restorations. There is no contraindication to applying SAP P_11_-4 to initial lesions on all surfaces, therefore one scenario estimated treatment of all (100%) initial lesions using SAP P_11_-4. Finally, we created one scenario (“NCTmix”) in which initial lesions were treated with the ideal noninvasive therapy with respect to the surface/location of the initial lesions in the mouth. The 24% of initial lesions occurring in pit and fissure surfaces in cohort data are treated with sealants; the 26% of initial lesions in facial surfaces of anterior teeth and premolars (cosmetic surfaces), where the SDF stain may not be acceptable to dental teams and patients, are treated with SAP P_11_-4; and the remaining 50% of initial lesions are treated with SDF. In the model, initial lesion diagnosis and management occur at baseline, and the treatment survival or failure is calculated at the end of each year for three years. The payor cost and clinic cost, gross revenue, net profit, and profit margin for each scenario, including treatment failure and repeat failure, are calculated using standard formulas (Appendix A).

**Figure 1.**
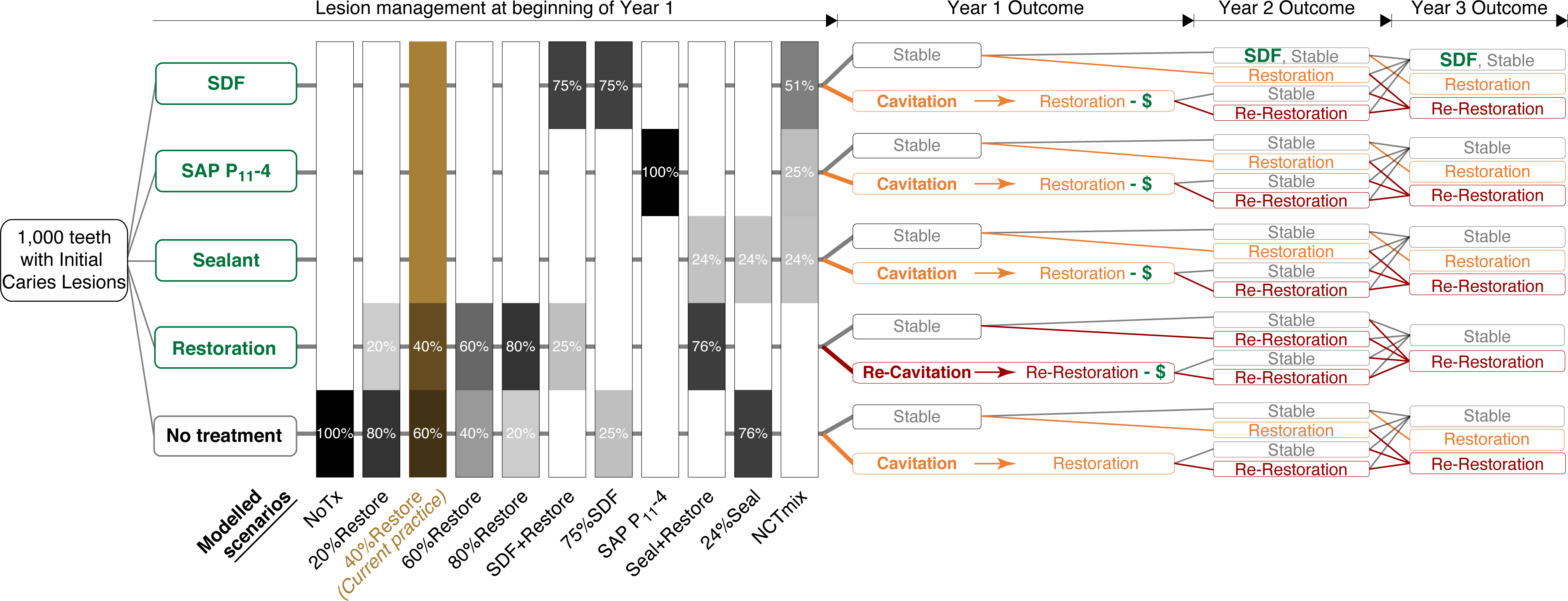
Initial caries lesion management model. The left side of the figure represents a table with columns corresponding to the scenarios and rows corresponding to each management method. The 11 modeled scenarios (vertical bars) are simulated by applying the relevant fraction of teeth into each year 1 management category (green and black at left). The current practice scenario model is highlighted in brown. Payor costs and clinic costs and net revenue are incorporated for the initial treatments (green) and any progression of caries lesions reaching cavitation and therefore indicating a restoration (orange) or re-restoration (crimson). Restoration from treatment failure within one year is reimbursed with the initial treatment fee removed (-$); restoration thereafter is reimbursed fully. Teeth without restoration each year are considered stable (grey) and have no costs.

### Sensitivity analysis

A Probabilistic Sensitivity Analysis (PSA) was conducted with ten thousand iterations to create more accurate estimates and to assess the reliability of results. Hourly wage ranges of the 10^th^ to 90^th^ percentile for dentists, hygienists, and dental assistants were used with an assumption of a triangle (linear) distribution as the input parameters of salary for the three clinician types vary based on years of experience and location. The failure ranges for each treatment or non-treatment were derived using an assumed beta distribution around the arithmetic mean failure rate, by considering the number of actual failures and observations. We modeled a uniform range of reimbursement for repeat application of SDF in the 2^nd^ and 3^rd^ years, and for the reimbursement of SAP P_11_-4. As the price of SAP P_11_-4 may change because it is a new product, we have used a uniform distribution with range of $27 to $33 for V2 (1-3 teeth) and $40 to $50 for V3 (4-6 teeth). In the PSA, the start sample of the scenarios was fixed to 1,000 ICLs at the beginning of the 3-year simulation, and random values were picked for each variable from a range and distribution as previously described and in Table 1, resulting in different costs and gross revenues for each scenario at the end of 3 years. This was repeated 10,000 times, and mean values with a range between 2.5 and 97.5 percentile are reported.

## Results

### Reimbursement estimate

SAP P_11_-4 reimbursement was estimated at $37 per tooth per the pricing analysis (Figure 2; Supplemental Table 2c). There is a range ($32 – $43) in which shifting from the current practice model to treating all teeth with SAP P_11_-4 increases payor savings and increases clinic net profit. Reimbursement at the midpoint increases clinic net profit by 41% and decreases payor costs by 10% compared to current practice. The midpoint represents a balanced reimbursement to incentivize both stakeholders to switch.

**Figure 2.**
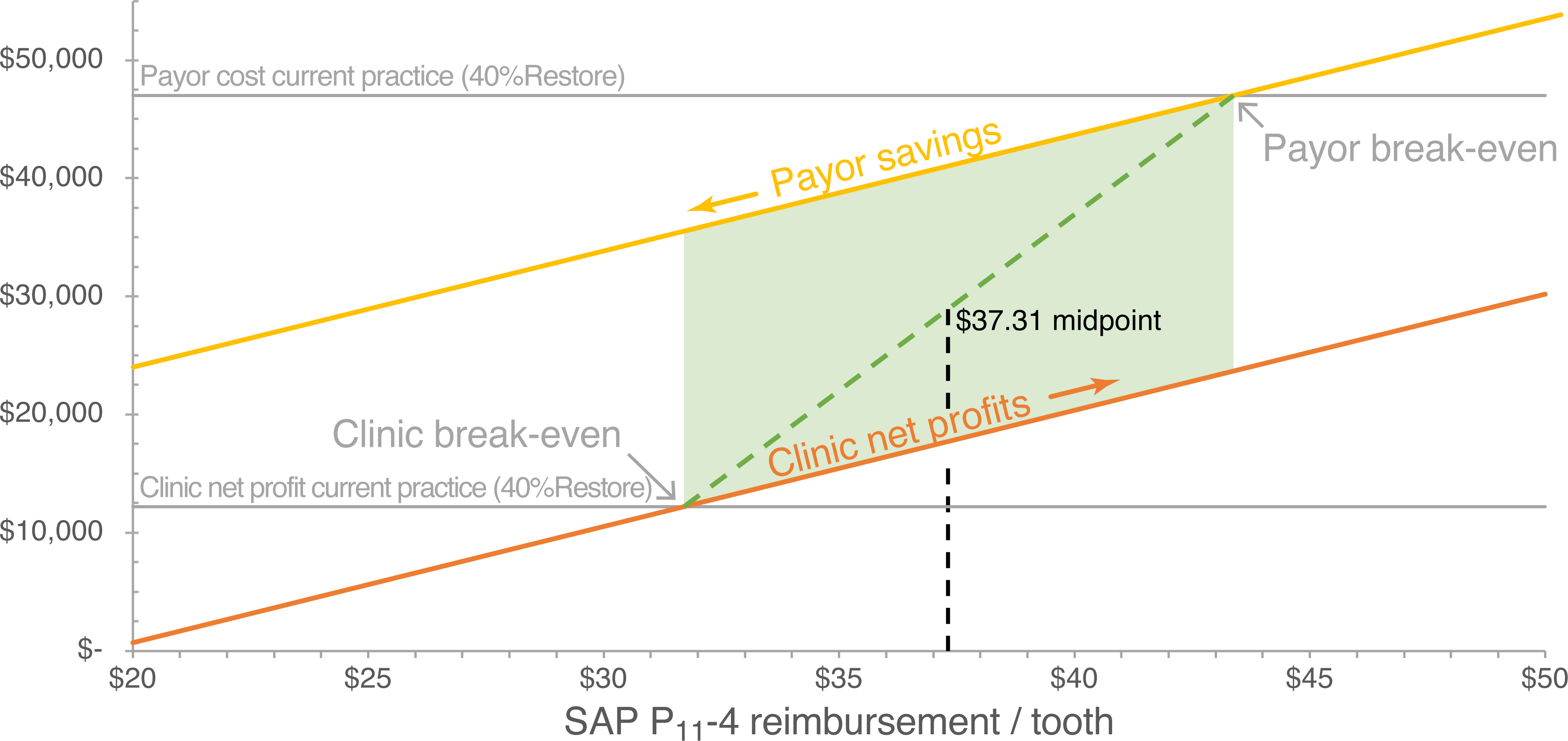
Modeling SAP P_11_-4 reimbursement to balance payor and clinic financial interests. Payor savings (yellow) and clinic net profit (orange) with respect to the current practice model of 40% restorations are shown for a range of per-tooth reimbursement amounts for SAP P_11_-4 treatment.

### Treatment Survival

Survival of treatments (i.e., without receiving restoration after baseline treatment) across three years is shown in Table 2. Drill-free tooth survival across three years for 1,000 initial lesions managed with each scenario are shown in Table 3.

### Savings and net profits

Cumulative payor costs, clinic costs and clinic net revenue from the deterministic analysis are shown broken out by year in Supplemental Figure 1 (numerical values in Supplemental Table 4), and in summary for all three years in Figure 3 (numerical values in Supplemental Table 5). All scenarios involving fewer restorations than the 40%Restore current practice model result in payor savings, in order from greatest savings to least: NoTx, 24%Seal, 75%SDF, NCTmix, 20%Restore, and SAP P_11_-4. SDF+Restore is highly similar in payor costs. Scenarios involving more restorations than the current practice model (60%Restore, Seal+Restore, and 80%Restore) result in higher payor costs.

**Figure 3.**
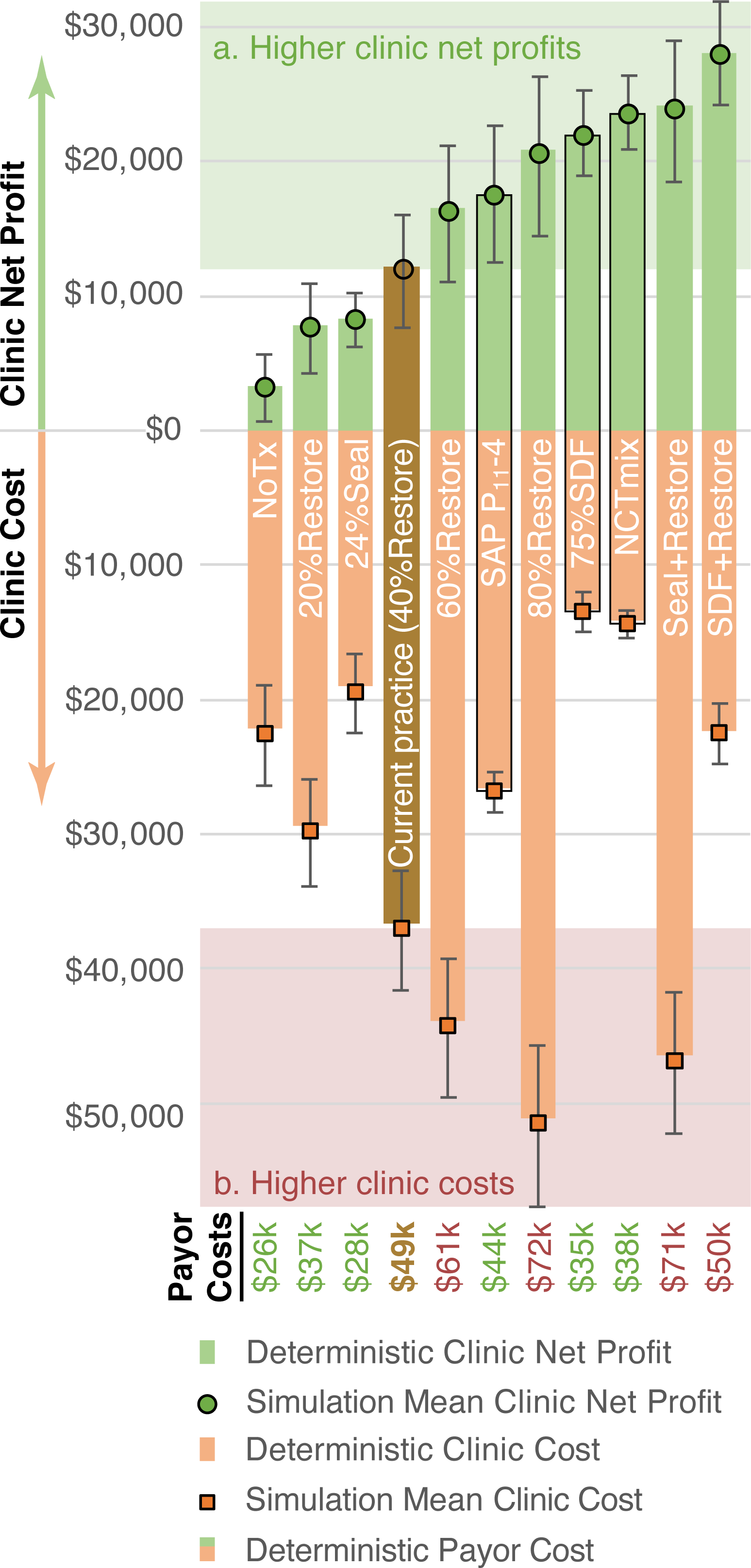
Increasing clinic net profit modified by clinic costs and payor costs. The clinic net profit (green) and costs (orange) at the end of 3 years are shown for each scenario, from deterministic (bars) and probabilistic (dots and 95% distributions) approaches. All deterministic data are within the range observed in the corresponding simulations. Payor costs are shown as the combined bar height and is written at bottom. From a payor financial perspective, all seven scenarios with lower payor costs are improvements on the current practice scenario (shown as brown bar). From a clinical financial perspective, scenarios ending in box “a” and not extending into box “b” are optimal. Basing clinical decisions purely on net profit would lead to further overtreatment; considering clinic costs identifies three scenarios as improvements (black outline) from current practice.

Clinic costs follow the same pattern as payor savings, with all scenarios involving fewer restorations than the 40% current practice model resulting in reduced clinic costs, and those involving more restorations than the current practice model resulting in greater clinic costs. All scenarios that treat more teeth than the current practice model (SDF+Restore, NCTmix, Seal+Restore, 75% SDF, SAP P_11_-4, 80%Restore, and 60% Restore) result in higher clinic net profits. All deterministic outcomes fall within the ranges observed in the probabilistic outcomes, as seen layered together in Figure 3.

All scenarios including SAP P_11_-4 and/or SDF without restorations meet all three success parameters of increasing payor savings, reducing clinic costs, and increasing clinic net profit: SAP P_11_-4, 75%SDF, and NCTmix. SDF+Restore meets the clinic benefit criteria but does not reduce payor costs.

### Immediate clinic net profit per patient visit

At a per-patient visit level, the increased net profit of noninvasive therapies is minimal or absent for one tooth but becomes substantial with multiple teeth treated (Supplemental Figure 2). Net profits for noninvasive therapies follow an economy of scale by number of teeth, due to the trivial additional time and marginal increase in material costs for application to additional teeth. Distribution of restorations across the mouth make a substantial impact on restorative dentistry due to the increased dentist and assistant time (e.g., time for local anesthesia, use rubber dam isolation; Table 1). In comparison, distribution across the mouth makes trivial impact on noninvasive therapy profitability.

### Clinic net profit per hour

Clinic net profit per chair time is higher than the current practice model for all scenarios except NoTx and 20%Restore (Figure 4). Treating the most teeth at baseline with the least invasive procedures corresponds to the highest net profit per chair hour, with the top four being NCTmix ($328/hour), SAP P_11_-4 ($256/hour), 75%SDF ($232/hour), and SDF+Restore ($190/hour; Figure 4A). The use of time for all personnel for each scenario is plotted against the corresponding clinic net profit in Figure 4B. Using hygienist time for SDF and/or SAP P_11_-4 scenarios saves considerable dentist and assistant time while increasing clinic net profits compared to traditional restorations.

**Figure 4A and Figure 4B.**
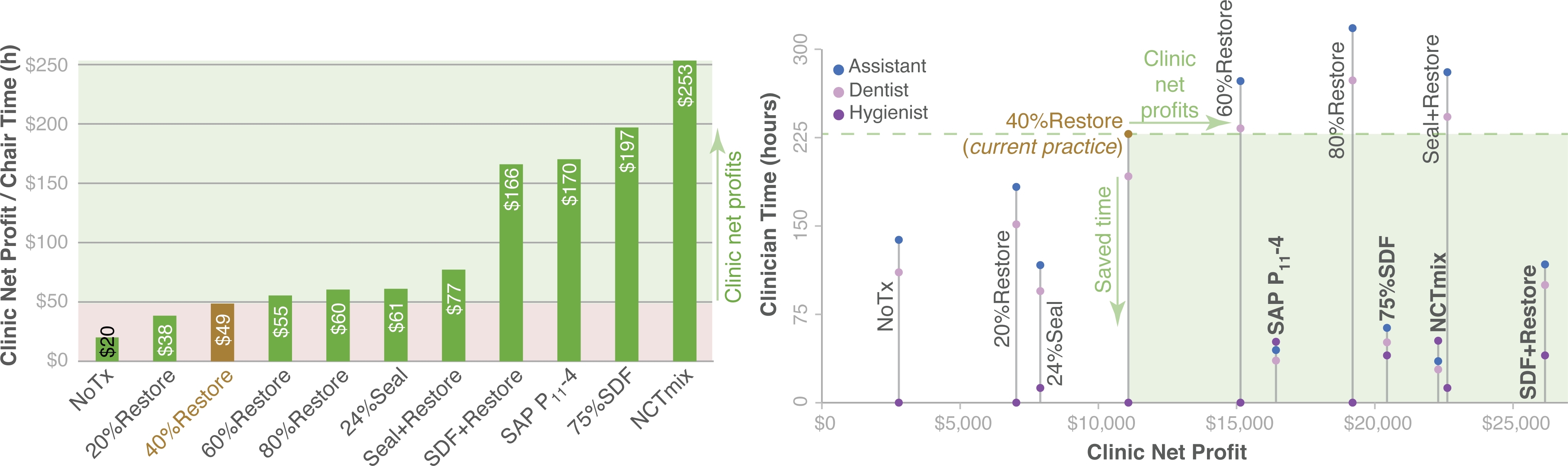
Clinic net profit per hour. Left |. **A**) Clinic net profit divided by the chair time throughout 3 years is shown for each scenario. Higher clinic net profit per chair time occurs when the corresponding bar height is higher than that of the current practice model (green box). All scenarios with non-invasive caries therapies are more profitable per chair time than all restorative-only scenarios. **Right | B**) Clinic net profit mapped against use of time from each clinician type: hygienist (purple), dentist (lilac), and assistant (blue). Times for each scenario are connected by a grey line. Bolded scenarios use the least amount of clinician time (often the most limited resource) while maximizing net profit.

### Financial and patient benefits

Payor savings, clinic profit margin, and drill-free tooth survival (patient benefits) are simultaneously improved for all non-restorative NCT scenarios (Figure 5). Restorative-focused scenarios have inverse relationships with payor savings and clinic profit margin. Restorative-focused scenarios with less than 40% restorations increase payor savings and drill-free tooth survival at the expense of decreased clinic profit margin. (Figure 5). The scenarios with the highest drill-free survival in order are NCTmix, SAP P_11_-4, 75%SDF, and 24%Seal (Supplemental Table 3).

**Figure 5A and Figure 5B.**
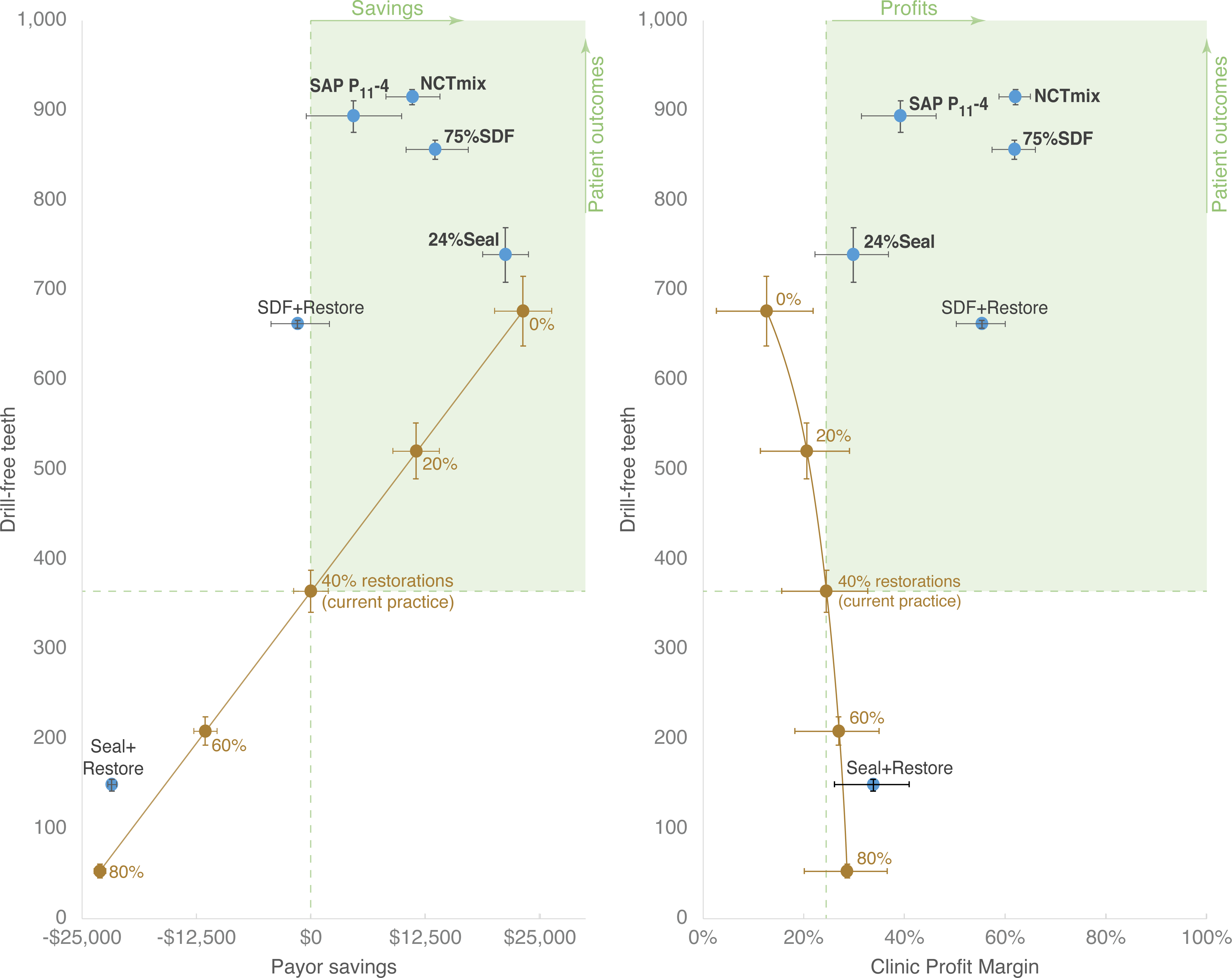
Comparison of payor savings and clinic profit margin to drill-free teeth. Left |. **A**) Payor savings and **Right | B)** Clinic profit margin for each initial caries lesion management scenario is plotted against teeth surviving without any caries progression to cavitation at the end of 3 years. Bolded scenarios improve patient outcomes, payor savings, and clinic profit margin. All nonrestorative scenarios with SDF and/or SAP P_11_-4 are superior for all stakeholders.

When considering scenarios with greater treatment survival (preventing the need for additional restorative treatments after baseline), scenarios including SAP P_11_-4 and/or SDF without restorations lead to reduced payor and clinic costs as the number of surviving treatments increase (Supplemental Figure 3; NCTmix, 75%SDF, and SAP P_11_-4). Non-SDF and non-SAP P_11_-4 scenarios that treat fewer teeth at baseline than the current practice model increase payor savings, but produce fewer surviving treatments and have lower clinic net profit (24%Seal, 20% Restore, and NoTx). Scenarios involving more restorations have higher payor costs and marginally higher clinic net profit (60%Restore, 80%Restore, and Seal+Restore). The scenarios with the highest clinic net profit in order are, SAP P_11_-4, SDF+Restore, 75%SDF, and NCTmix.

## Discussion

Our study findings suggest that shifting from the current practice model (40%Restore) to non-restoratively treating initial caries lesions with SDF or SAP P_11_-4 (or a mix thereof) results in payor savings, higher clinic net profit, and superior outcomes for patients. Additionally, all scenarios that treated more teeth at baseline than the current practice model resulted in more treatments surviving after 3 years. Longer-term payor savings may be even greater than our modeling shows, as costs accumulate over time due to the progression of initial lesions to cavitation and restoration.

When considering the chair and clinician time needed to achieve increased net profits, the noninvasive approaches NCTmix, SAP P_11_-4, 75%SDF, and SDF+Restore demonstrate clear superiority. The NCTmix net profit per chair time is six times that of the current practice model. SAP P_11_-4 and 75%SDF resulted in a net profit per chair time over four times the current practice model. When considering clinic costs and clinician time needed to achieve clinic net profit, four scenarios emerge as superior: SDF+Restore, NCTmix, 75%SDF, and SAP P_11_-4. By utilizing noninvasive therapies, dental assistants and hygienists can work to the highest level of their scope of practice, freeing up dentists to deliver care that requires advanced training. More complex procedures tend to have higher net profit per time, creating opportunities for higher profits (not represented in this work). Shifting production from managing initial lesions with restorations to performing higher net profit procedures amplifies the financial benefits of managing initial lesions with SDF and SAP P_11_-4.

Noninvasive therapies offer a new way to achieve improved treatment outcomes for patients while financially incentivizing payors and clinics. They follow an economy of scale by teeth treated per patient visit, so that two or more teeth treated per patient visit becomes increasingly more profitable. When considering chair time, the mix of noninvasive therapies provides the highest clinic net profit, with SAP P_11_-4 and SDF emerging as much more profitable than other scenarios. The drivers of payor savings and clinic net profit for SDF and/or SAP P_11_-4 scenarios are minimal treatment times, combined lower cost of material and clinician labor (compared to traditional restorations), and the applicability to most or all tooth surfaces. Improvements in patient outcomes that are financially rewarding for clinics and payors are hallmarks of value-based care. As the oral health care system makes meaningful, albeit gradual, progress toward implementing value-based care, “there will be greater financial incentives [for oral health care clinics]…to avoid invasive surgical interventions.”^21^ Noninvasive therapies are a critically important tool in the value-based toolbox within oral health care.

Treatment of initial lesions with traditional restorations is extremely common, as 94% of US dentists report restoring initial lesions^5^ while only 11%-12% of control group initial lesions progress.^6, 22^ This disconnect may be due to the ubiquitous misunderstanding that radiographic appearance of demineralization reaching the dentin necessarily indicates cavitation.^5, 23, 24^ However, international guidelines recommend against restoring initial lesions as these lesions can be healed, or arrested, through remineralization.^2–4^ Although restorations have treatment failure rates similar to noninvasive therapies, when noninvasive therapies fail the negative result is a restoration, whereas failed restorations require deeper and more invasive procedures. Noninvasive therapies provide a treatment option that empowers dental teams to heal the first signs of disease.

Sealants are a highly efficient and successful method of treating initial lesions and contribute to the superiority of the NCTmix in the current analysis. The ADA recommends sealants for the management of initial lesions.^3, 13, 14^ Additionally, the World Health Organization (WHO) recently added glass ionomer, modeled here as the sealant material, to their Lists of Essential Medicines for Children and for Adults in recognition of its preventive and therapeutic value.^25–27^ The optimal result of the NCTmix scenario demonstrates that use of sealants to manage initial lesions is ideal when supplemented with other noninvasive therapies (e.g., SDF or SAP P_11_-4), rather than being used with other restorations or alone. Since diagnosis is not widely implemented in American dentistry, payors and clinics may choose to use a CDT code that indicates sealants placed over initial dental caries lesions for tracking purposes (e.g. D1352 – preventive resin restoration, or D2940 – protective restoration).

In the short time since it has been used in the US for caries arrest, SDF has already been impactful, for example by lowering the use of general anesthesia in young children by 70% in a large Medicaid-focused network of community clinics.^28^ SDF is the focus of the ADA evidence-based guideline on NonRestorative Caries Treatment.^3^ The WHO acknowledged SDF’s potential to equitably decrease suffering from caries by adding it to its Lists of Essential Medicines (for both children and adults).^3, 24, 26^ The simplicity of applying SDF was also recently recognized by the creation of a medical procedure code.^26, 29^ However, current evidence primarily focuses on use for moderate to advanced caries lesions rather than initial lesions.^3, 10, 30^ The results of this study provide evidence of the cost-effectiveness of SDF for treatment of initial lesions.

SAP P_11_-4 is a relatively new drug that provides a therapeutic effect to rebuild enamel hydroxyapatite. Unlike SDF, its lack of staining and neutral flavor positions it to be used throughout the mouth, and particularly for permanent anterior teeth.^31^ It is not used to seal in caries, and therefore may be more accepted by dentists than sealants.^31, 32^ For dental clinics, the profit margin for SAP P_11_-4 is superior, because it takes less clinician time to perform the application than to place restorative fillings.

While not a direct focus of this analysis, noninvasive therapies are likely to be more beneficial for, and favored by, patients over traditional dental treatments for initial lesions. Dental fear and anxiety are well-documented barriers to receiving regular oral health care;^33–35^, and should be reduced given that noninvasive therapies are pain, needle, and drill-free. Since noninvasive therapies can be easily delivered by non-dentist professionals within and outside traditional dental care settings (e.g., schools, communities), they should improve patient access to oral health care. Finally, since noninvasive therapies stop and reverse disease progression, they help patients keep their teeth longer and avoid expensive restorative care resulting from recurrent caries lesions.^36^

This study includes some limitations. The clinical outcome data on which the analysis depends cannot be representative of all populations, and likely represent less disease progression than in the population. This case study of Alabama Medicaid may not represent challenging dynamics in other geographic areas, though any can be similarly modeled. Medicaid fee schedules usually represent lower fees than commercial insurance reimbursements. The scenarios are meant to demonstrate practical options in clinical practice but cannot represent all constellations of mixes. Real-life challenges will inevitably deviate from the outcomes of this theoretical model, particularly during the change from a restorative to a non-invasive paradigm; however, these data predict strong financial benefits after a transition time.

## Conclusion

This work finds that the use of noninvasive therapies for initial lesions increases payor savings, is efficient and profitable for clinics, and is beneficial for patients in the modeled scenarios. The key component of the economic success of noninvasive therapies is to empower non-dentist clinicians to treat dental caries. Accordingly, to replicate the success of this noninvasive paradigm cost model 1) policymakers should support non-dentist clinicians to manage initial lesions; 2) payors should invest in creating value-based reimbursements, and 3) clinics should adopt new procedures with consistent change management. The time for these techniques has come.

## Figure Legends

**Supplemental Figure 1: Deterministic clinic net profit, clinic costs, and payor costs**. The clinic net profits (green), clinic costs (orange), and payor costs (total) at the end of each year for 1,000 initial caries lesions by each management scenario. Payor savings (yellow and brown dashed line) occurs when the year 3 total bar height is less than that of the current practice model (brown outline). Increased clinic net profit (dashed green and brown line) occurs when the year 3 green bar is higher than that of the current practice model (brown outline). All options that include SDF and/or SAP P_11_-4 increase clinic net profit while also producing payor savings.

**Supplemental Figure 2. Per patient visit immediate clinic net profit by number of teeth and anatomical areas**, with immediate costs only. Distribution of the teeth across anatomic areas (A) of the mouth (e.g., quadrants) is much less impactful on noninvasive caries therapy net profit versus restoration net profit because there is no need for local anesthesia or rubber dams. Treating more teeth (T) per patient in one visit is more profitable for noninvasive caries therapies regardless of anatomic distribution; all caries lesions can be managed in one visit.

**Supplemental Figure 3A and Figure 3B. Comparison of payor savings and clinic profit margin to treatment survival. Left | A)** Payor savings and **Right | B)** Clinic profit margin for each initial caries lesion management scenario is plotted against treatments surviving without caries progression to cavitation at the end of 3 years. Bolded scenarios improve patient outcomes, payor savings, and clinic profit margin. All nonrestorative scenarios with SDF and/or SAP P_11_-4 are superior for all stakeholders.

## Data Availability

All data produced in the present study are available upon reasonable request to the authors.

## Acknowledgements

Special thanks to the clinical innovation, operations, and data analytics teams of Advantage Dental+. Thanks to Dr. Leah Price for guiding the beginnings of this work. Thanks to our colleagues of CareQuest Institute for Oral Health for support and critical feedback, particularly Kelly Schroeder and Caroline McLeod. Thanks to Jean-Francois Blouvac of vVARDIS and to Steve Pardue of Elevate Oral Care for insightful conversations on the dynamics of costs and benefits in oral health product innovation.LJLJ

## Contributions

Conceptualization (JHK, LK, CD, KZ, JW, SS); design & investigation (JHK, LK, SS, JW); analysis (JHK, LK, LH, CD, SS, JW, KZ, MF); writing (JHK, LK, LH, SS); and editing (JHK, LK, LH, CD, SS, JW, KZ, MF).

## Appendix A: Key Terms Defined

|  |  |
| --- | --- |
| Payor cost(s) | The amount of money that the payor pays the clinic for services provided to their covered members. In this model, this is the same as the clinic's gross revenue. |
| Payor savings | The financial benefit realized from reducing the payor's payments (payor costs). In this financial model payor savings occur anytime a scenario costs less than the current practice scenario – represented by the 40% restoration scenario, for services provided to manage 1000 initial lesions. |
| Clinic gross revenue | Total revenue generated from clinic operations or services provided. |
| Clinic cost(s) | The amount of money that the clinic pays to provide services to their patients. This includes expenses such as labor, overhead, and material costs. |
| Clinic net profit(s) | The amount of revenue remaining after all clinic costs are deducted from gross revenue.<br>Clinic net profit = Clinic gross revenue – clinic costs |
| Clinic profit margin | A measure of clinic profitability that is expressed as a percentage. This measure displays the portion of gross revenue that the clinic keeps after accounting for clinic costs.<br>Clinic profit margin = Clinic net profit / clinic gross revenue |
| Payor Financial benefit | In this work, a payor financial benefit is defined as payor costs (fees paid out to the clinic for services provided in each scenario) being less than the payor costs of current practice. This could also be reported as increased payor savings compared to current practice. |
| Clinic Financial benefit | In this work clinic financial benefits include increased clinic net profit and increased profit margin. These benefits are often realized through decreased clinic costs, increased time efficiency, and increased gross revenue. |
| Patient benefit | In this work, patient benefits include improved treatment survival rates which reduced the need for additional restorative visits after baseline treatment. Patient benefits can also include the number of teeth that survive without ever needing restorative treatment or drill-free teeth. Other indirect patient benefits include shorter appointment duration, and increased access to care. |

