## Supplemental Table 1 for "SDF, SAP P_11_-4, and GI Sealants for Managing Initial Caries Lesions Produce Clinic and Payor Savings in Financial Impact Model" (from another version of this preprint: version 1, 2023-01-31)

**Supplemental Table 1:** Probability of combination between one to six teeth and one to four anatomical areas and the number of teeth that can be in each combination out of 1,000 teeth.

| Number of teeth | Quadrant | Probability of the combination |
| --- | --- | --- |
| 1 | 1 | 24.77% |
| 2 | 1 | 8.15% |
| 2 | 2 | 12.14% |
| 3 | 1 | 1.98% |
| 3 | 2 | 7.80% |
| 3 | 3 | 4.64% |
| 4 | 1 | 0.71% |
| 4 | 2 | 4.08% |
| 4 | 3 | 5.23% |
| 4 | 4 | 1.47% |
| 5 | 1 | 0.32% |
| 5 | 2 | 1.51% |
| 5 | 3 | 3.84% |
| 5 | 4 | 2.11% |
| 6 | 1 | 0.43% |
| 6 | 2 | 0.96% |
| 6 | 3 | 4.98% |
| 6 | 4 | 14.88% |
| Total |  | 1 |
