## Supplemental Table 2 for "SDF, SAP P_11_-4, and GI Sealants for Managing Initial Caries Lesions Produce Clinic and Payor Savings in Financial Impact Model" (from another version of this preprint: version 1, 2023-01-31)

**Supplemental Table 2:** Deterministic estimates of cost to clinician, net profit, and cost to payor for each year compared across 11 scenarios. Data also represented in Supplemental Figure 1.

|  | Year 1 | | Year 2 |  | Year 3 |  |
| --- | --- | --- | --- | --- | --- | --- |
| Scenarios | Cost to Clinic | Net Profit | Cost to Clinic | Net Profit | Cost to Clinic (Less the better) | Net Profit |
| NoTx | $7,929 | $1,302 | $15,033 | $2,316 | $ 21,566 | $3,368 |
| 20%Restore | $16,726 | $5,979 | $22,647 | $6,928 | $28,145 | $7,849 |
| 40%Restore | $25,522 | $10,656 | $30,261 | $11,455 | $34,725 | $12,245 |
| 60%Restore | $34,319 | $15,334 | $37,875 | $15,982 | $41,304 | $16,641 |
| 80%Restore | $43,115 | $20,011 | $45,489 | $20,509 | $47,883 | $21,037 |
| 75%SDF | $4,588 | $12,709 | $8,102 | $19,036 | $11,155 | $22,448 |
| Seal+Restore | $40,745 | $23,642 | $41,825 | $23,934 | $43,037 | $24,263 |
| Curodont | $20,698 | $17,496 | $22,050 | $17,708 | $23,398 | $17,921 |
| 25%Seal | $7,450 | $5,938 | $13,007 | $6,799 | $18,361 | $8,326 |
| Seal+Restore | $40,745 | $23,642 | $41,825 | $23,934 | $43,037 | $24,263 |
| NCTmix | $8,369 | $17,687 | $10,065 | $21,863 | $11,488 | $23,913 |
