## Supplemental Table 3 for "SDF, SAP P_11_-4, and GI Sealants for Managing Initial Caries Lesions Produce Clinic and Payor Savings in Financial Impact Model" (from another version of this preprint: version 1, 2023-01-31)

**Supplemental Table 3:** Probabilistic estimates of cost to clinician, net profit, and cost to payor for each year compared across 11 scenarios. Data also represented in Figure 3.

| Scenarios | Payor cost  (Clinic revenue) | | | Clinic cost | | | Clinic profit | | |
| --- | --- | --- | --- | --- | --- | --- | --- | --- | --- |
|  | Mean | 2.5^th^% | 97.5^th^% | Mean | 2.5^th^% | 97.5^th^% | Mean | 2.5^th^% | 97.5^th^% |
| NoTx | $25,220 | $22,184 | $28,270 | $21,936 | $18,452 | $25,696 | $5,648 | $800 | $2,484 |
| 20%Restore | $36,224 | $33,783 | $38,679 | $28,507 | $24,833 | $32,475 | $10,804 | $4,416 | $3,301 |
| 40%Restore | $47,142 | $45,312 | $48,988 | $35,077 | $31,018 | $39,479 | $15,892 | $7,949 | $4,116 |
| 60%Restore | $58,061 | $56,837 | $59,289 | $41,647 | $36,944 | $46,688 | $21,016 | $11,474 | $4,939 |
| 80%Restore | $68,979 | $68,366 | $69,597 | $48,218 | $42,818 | $54,046 | $26,139 | $15,000 | $5,762 |
| SDF+Restore | $47,468 | $44,481 | $51,044 | $19,390 | $17,529 | $21,409 | $31,858 | $24,528 | $3,549 |
| 75%SDF | $33,702 | $30,480 | $37,464 | $11,286 | $10,003 | $12,651 | $25,713 | $19,513 | $2,903 |
| Curodont | $41,850 | $35,926 | $47,843 | $23,912 | $22,323 | $25,859 | $23,476 | $12,422 | $5,516 |
| Seal+Restore | $67,446 | $67,137 | $67,829 | $43,430 | $38,711 | $48,478 | $28,726 | $18,962 | $5,054 |
| 24%Seal | $27,059 | $24,613 | $29,553 | $18,791 | $16,068 | $21,760 | $10,187 | $6,264 | $2,004 |
| NCTmix | $35,692 | $32,719 | $38,905 | $11,786 | $10,932 | $12,691 | $26,802 | $21,174 | $2,731 |
