## Supplementary figures and images for "SDF, SAP P_11_-4, and GI Sealants for Managing Initial Caries Lesions Produce Clinic and Payor Savings in Financial Impact Model" (from another version of this preprint: version 1, 2023-01-31)

### Supplemental Figure 1

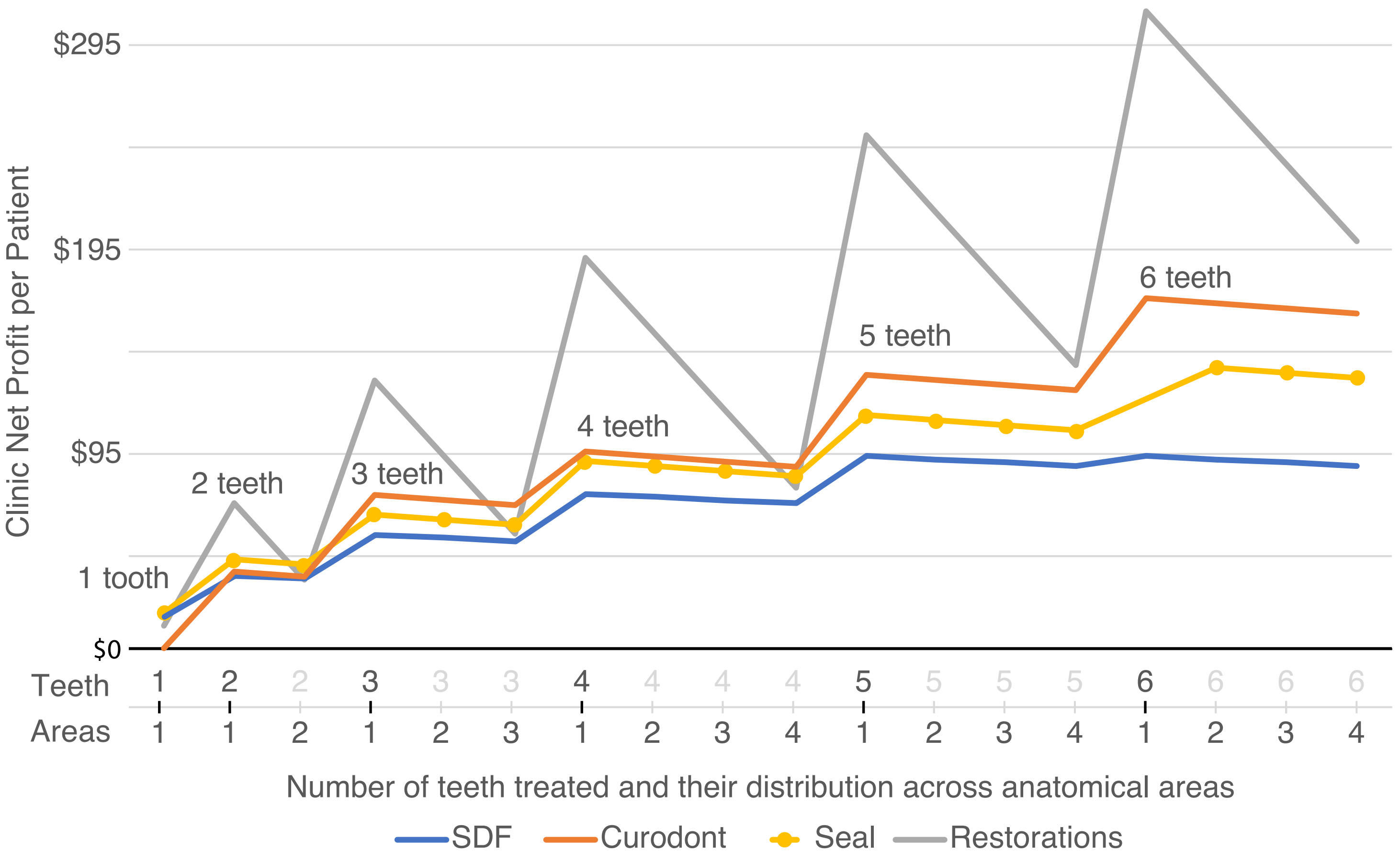

### Supplemental Figure 2

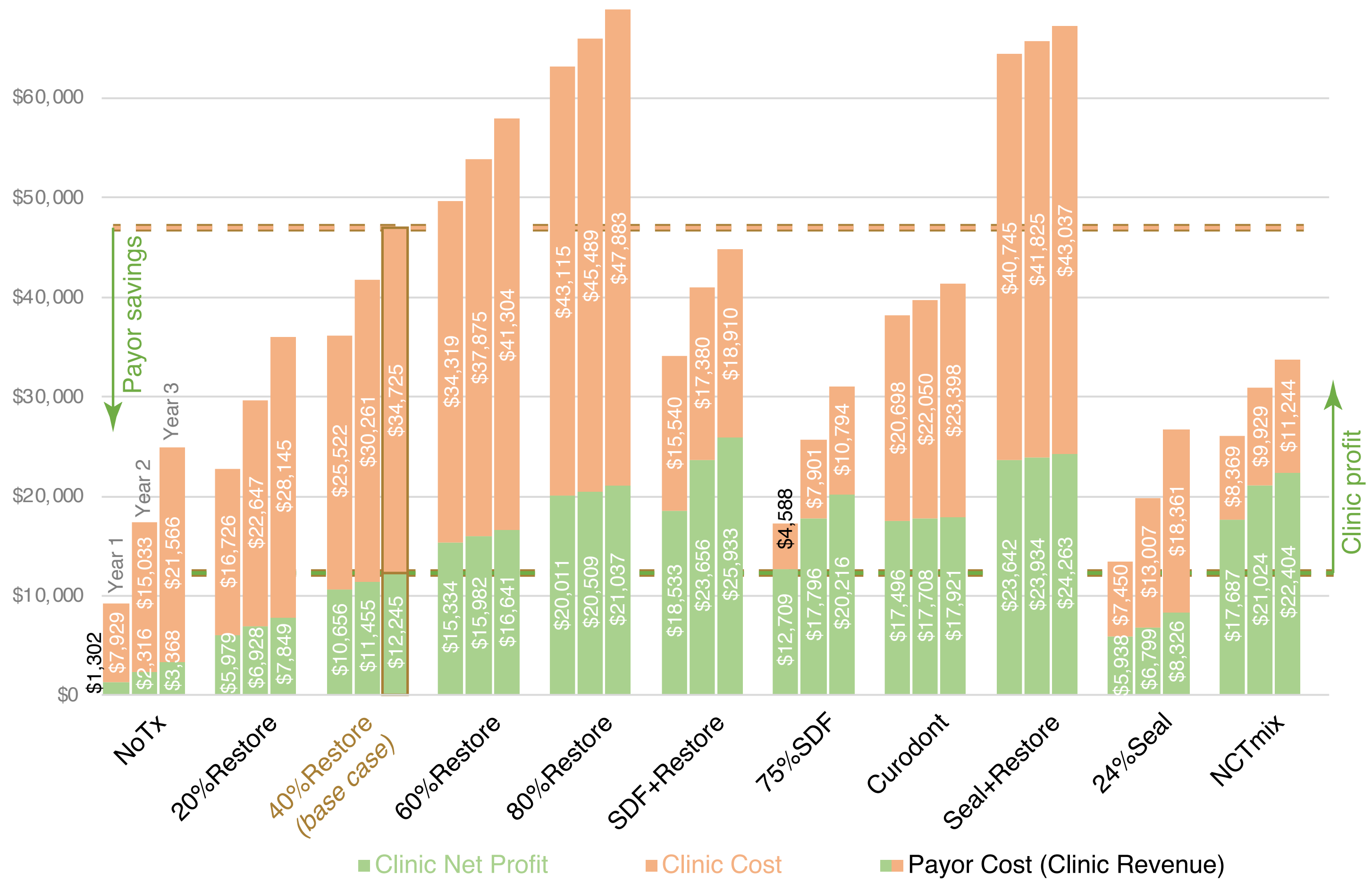

### Supplemental Figure 3

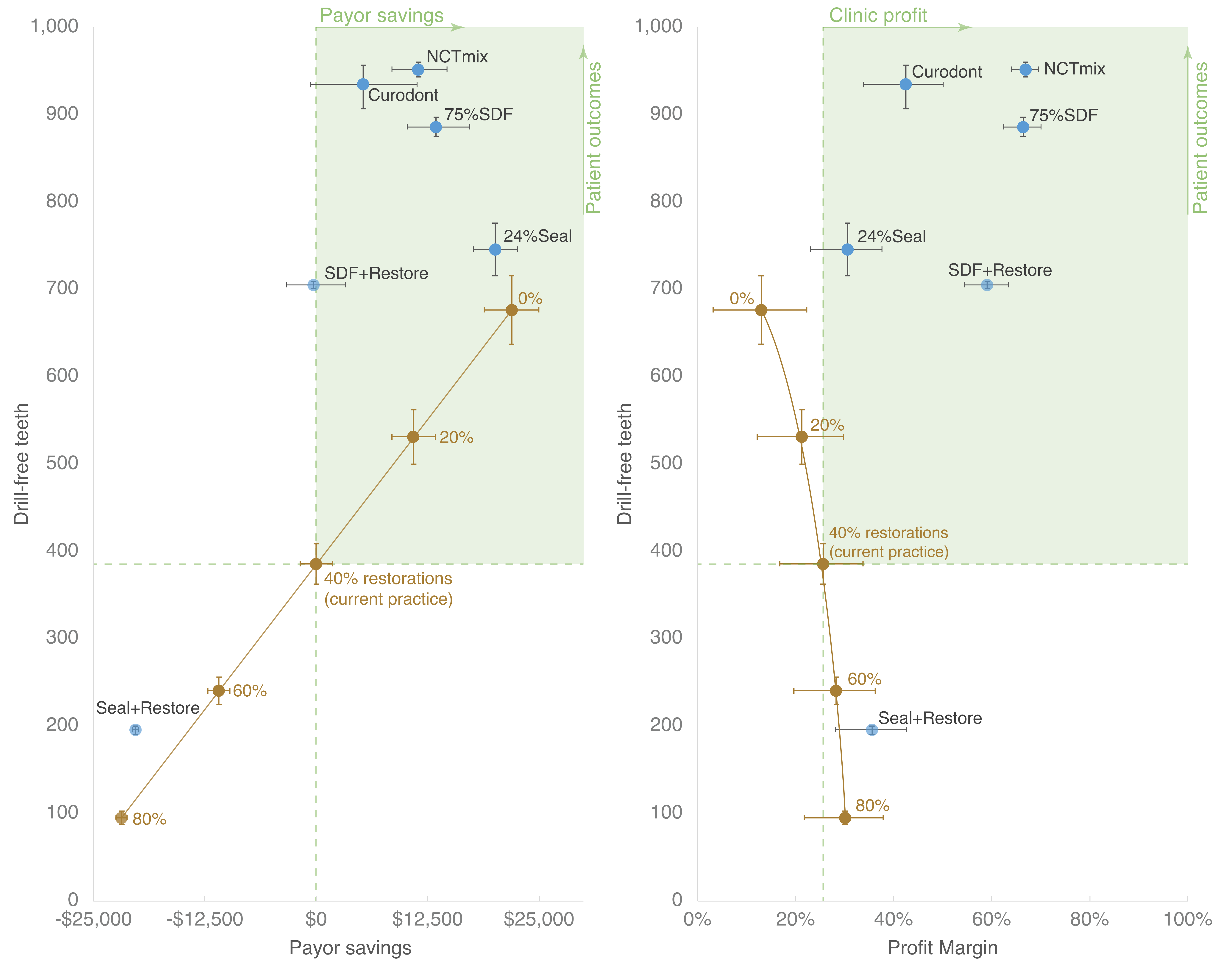
